# Development and Validation of a Pictorial Cue Database for Cannabis Cue Reactivity Studies: Insights from Behavioral and Neural Investigations

**DOI:** 10.1101/2024.07.26.24310880

**Authors:** Zahra Hamidein, Neda Mohammad, Parnian Rafei, Mohsen Ebrahimi, Hamed Ekhtiari, Peyman Ghobadi-Azbari, Tara Rezapour

**Author notes:** **Corresponding authors:** 1. Tara Rezapour, Department of Cognitive Psychology, Institute for Cognitive Science Studies (ICSS), Tehran, Iran; 2. Peyman Ghobadi-Azbari, Iranian National Center for Addiction Studies, Tehran University of Medical Sciences, Tehran, Iran.

## Abstract

**Introduction:** Craving, a potent driving force behind drug-seeking and consumption behaviors, represents a dynamic emotional-motivational response primarily elicited by drug-related cues. In laboratory settings, the drug cue reactivity (DCR) paradigm is frequently employed to evoke craving and investigate the neural and behavioral responses to drug cues. This study adopts functional magnetic resonance imaging (fMRI) alongside behavioral assessments to establish a collection of validated pictorial cues encompassing both cannabis and neutral images.

**Methods:** 110 cannabis-related images were selected across cannabis flowers and powder, cannabis use methods, and paraphernalia categories. Participants with a history of cannabis use were then asked to assess the selected images for craving, valence, and arousal using both the visual analog scale and the self-assessment Manikin. Using fMRI, the neural mechanisms underlying cannabis cue-reactivity were investigated at the whole-brain level and within Brainnetome atlas areas in a subgroup of 31 cannabis users.

**Results:** The selected cannabis-related images (n = 110) received significantly higher craving (t = 6.56; p<0.001) and arousal (t = 17.46; p<0.001) ratings compared to the neutral ones (n = 30). Fifty regular cannabis users (19.9 ± 4.8 years; 10 females and 40 males) with at least a one-year history of use were included. Investigating blood oxygenation level-dependent (BOLD) responses to cannabis compared with neutral cues yielded significant activations in the inferior/medial frontal gyrus, fusiform gyrus, parahippocampal gyrus, orbital gyrus, postcentral gyrus, insula, precuneus, superior/middle temporal gyrus, and cerebellar tonsil.

**Conclusion:** This study provides a resource of ecologically validated cannabis-related images useful for studies applying DCR as interventions or assessments for cannabis users.

## 1. Introduction

The drug cue reactivity (DCR) paradigm is commonly used in experimental studies for both assessments and interventions (Ekhtiari et al., 2019). A "Cue" refers to a stimulus containing drug-related features presented through various sensory modalities such as visual, auditory, audiovisual, tactile, olfactory, or gustatory stimuli, which induce emotional responses in individuals with substance use disorders (SUDs). Craving, as an emotional response to drug-related conditioned cues, is experienced by individuals with various forms of SUDs (Ekhtiari et al., 2016), including cannabis use disorder (CUD) (Sherman et al., 2018). Studies indicate that DCR can serve as the core for laboratory-based assessments of treatment efficacy for individuals with SUDs. Previous research has explored the role of DCR as an intervention within exposure therapy (Goldstein et al., 2007) and memory reconsolidation paradigms (Ekhtiari et al., 2019). Cue exposure has been shown to elicit reward-related neural activation (Cousijn et al., 2013; Karoly et al., 2019), subsequently increasing subjective craving (Bonson et al., 2002; Ekhtiari et al., 2016; Vollstädt-Klein et al., 2011, Johnson et al., 2002).

Given the importance of cue exposure, several studies have validated visual cues through databases (Ekhtiari et al., 2019; Macatee et al., 2021). The Normative Appetitive Picture System (NAPS) was the first published database specifically designed for appetitive images, including 18 alcohol, 6 cigarettes, 12 food, and 12 non-alcoholic beverage-related images (Stritzke et al., 2004). Similarly, Billieux and colleagues validated alcohol-related images by asking participants to rate 60 alcohol-related images for valence, arousal, and dominance (Billieux et al., 2011). Another study provided a validated database of pictorial cues for methamphetamine and opioids (Ekhtiari et al., 2019), which included 120 images for each substance rated by participants with a history of use. They also added 120 neutral images matched for their content (objects, hands, faces, and actions) with drug-related images to increase the potential for this database to be used in experimental DCR tasks (Ekhtiari et al., 2019). Additionally, Mactee and colleagues recently developed a database comprising 280 cannabis-related images across four cannabis paraphernalia categories (bowl, bong, blunt/joint, vaporizer), rated by regular cannabis users varying in primary cannabis use method. They also rated 80 neutral images matched to the cannabis images based on important confounding elements and characteristics (e.g., presence of human hands and faces) (Macatee et al., 2021).

DCR tasks utilized in functional magnetic resonance imaging (fMRI) studies represent an essential step toward integrating functional neuroimaging into clinical practice in addiction medicine (Ekhtiari et al., 2019). Cue-reactivity reflects increased motivational processing underlying continued substance use and relapse (Wang et al., 2022). SUDs are associated with greater cue reactivity in brain regions such as the orbitofrontal cortex, anterior cingulate cortex, striatum, ventral tegmental area, and amygdala (Ekhtiari et al., 2016). Several fMRI studies have examined brain function in cannabis users exposed to cannabis vs neutral stimuli during cue-reactivity tasks (Karoly et al., 2019; Sehl et al., 2021). Despite methodological heterogeneity, these studies consistently demonstrate significant activations in response to cannabis stimuli, including in the amygdala, parietal, striatum, and prefrontal cortex (Cousijn et al., 2013; Karoly et al., 2019).

As cannabis use continues to rise globally, there is an increasing need for the development of therapeutic interventions and assessments tasks within cue reactivity paradigms. However, existing cue databases have several gaps, including a lack of multiple categorizations (e.g., combining elements of actions and paraphernalia) and the use of a combination of behavioral (subjective rating) and neural methods to validate the cues. To address this gap, the present study utilized fMRI and behavioral data to provide a set of validated pictorial cues for cannabis and neutral images in a sample of regular cannabis users. The images were selected from cannabis alone, cannabis use methods, and three cannabis paraphernalia categories (blunt/joint, pipe/bowl, and bong). Cannabis users also rated 30 neutral images matched to the selected cannabis-related images based on important features, including the presence of hands and faces.

## 2. Methods

The present study consisted of three phases (Figure 1): (a) pilot phase (cannabis cue validation), (b) behavioral phase (cue validation), and (c) neural phase (cannabis cue reactivity), which are described below, respectively.

**Figure 1.**
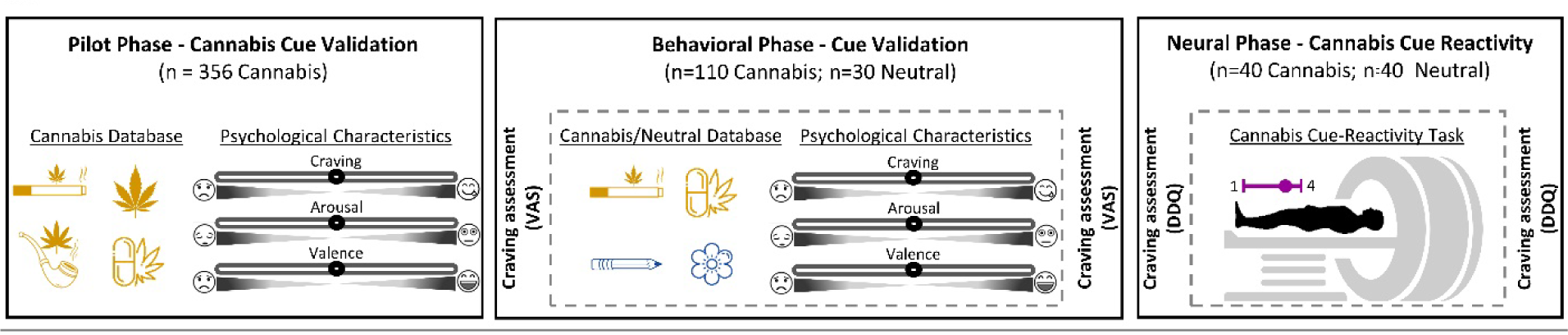
Overview of the experimental procedures and cannabis cue-reactivity paradigm. Experimental procedure includes three phases: pilot phase (cannabis cue validation), behavioral phase (cue validation), and neural phase (cannabis cue reactivity). In the cannabis cue validation phase (pilot phase), participants (n = 10) saw cannabis cues (n = 356) and rated the craving, arousal, and valence induced by each image. In the behavioral phase (cue validation), participants (n = 50) saw neutral cues and cannabis cues selected from the pilot study and rated the craving, arousal, and valence induced by each image. Immediately before and after the behavioral phase, participants rated their self-reported craving. In the neural phase (cannabis cue reactivity), participants (n = 31) underwent an MR scan with the cannabis cue-reactivity task. Immediately before and after the cue-reactivity task, participants completed the Desires for Drug Questionnaire (DDQ).

### 2.1. Pilot Phase: Cannabis Cue Validation

The pilot study was conducted face-to-face to select a set of cannabis-related images. A sample of 10 regular cannabis users participated, and their craving induced by each image was assessed. Images were displayed on a 17-inch LCD monitor positioned approximately 70 cm away, using a laptop (images were presented by Photo Viewer for Win 10 1.0 for Windows).

### 2.2. Behavioral Phase: Cue Validation

The main study utilized the images chosen from the pilot phase. These images were presented to two subgroups of cannabis users: one in an online behavioral phase (n = 50) and another in a neural phase employing an fMRI cannabis cue-reactivity task (n = 31).

### 2.3. Neural Phase: Cannabis Cue Reactivity

The neural phase involved the use of functional magnetic resonance imaging (fMRI) during a cannabis cue-reactivity task. This phase aimed to investigate neural responses to cannabis-related cues among regular cannabis users.

### 2.4. Participants

The inclusion criteria for the three phases were as follows: (1) fluency in Farsi, (2) age between 18 and 40 years, and (3) cannabis use at least twice per week over the past year. Participants were recruited via social media platforms such as Twitter and Instagram. Volunteers who met the inclusion criteria were selected and screened for eligibility. Additional inclusion criteria for participants in the fMRI study included: (1) abstinence from other substances and psychiatric prescription medication, (2) abstinence from cannabis for at least 12 hours prior to the scanning sessions (controlled by oral fluid testing), and (3) eligibility for MRI scanning.

For the pilot study, participants were invited to the laboratory to perform the DCR task and rate the images. For the online behavioral study, those who met the criteria received an online link containing questionnaires and a consent form prior to starting the cue validation task. All three phases of the study were conducted on the online Gorilla platform (https://gorilla.sc/).

Participants selected for the fMRI study were invited to the National Brain Mapping Lab, Tehran, Iran (https://www.nbml.ir) for imaging sessions. The study was approved by the Ethics Committee of the Iran University of Medical Sciences (Approval ID IR.IUMS.REC.1400.510), and all participants provided written informed consent before participation.

### 2.5. Materials

- *Demographic data:* Participants were asked to fill out a questionnaire providing information about their: age, gender, and education level, as well as history of cannabis use (i.e., duration of regular cannabis use and frequency of use per week). Additionally, participants confirmed their abstinence from other drugs and psychiatric prescription medication.
- *Cannabis-related images:* During the pilot study, 356 cannabis-related images were selected from two databases validated by Mactee (Macatee et al., 2021) and Karoly (Karoly et al., 2019). Out of these, 20 images depicted cannabis plant and powder, while the remaining images portrayed specific methods of use and paraphernalia categories (i.e., vaporing, smoking). In face-to-face sessions, the selected images were presented to 10 participants who rated affective measures including craving, arousal, and valence for each image. From this pilot study, 110 images with the highest mean craving scores and the most compatibility with Iranian cultural norms (without any sexual content) were selected for the online behavioral study. These images were categorized into Cannabis alone (subdivisions: cannabis flower and cannabis powder), Cannabis-related paraphernalia objects (subdivisions: blunt/joint, pipe/bowl, and bong), Cannabis-related paraphernalia with hands (subdivisions: blunt/joint with hand, pipe/bowl with hand, and bong with hand), and Cannabis-related paraphernalia activities with faces (subdivisions: blunt/joint with face, pipe/bowl with face, bong with face). Additionally, 30 toothbrush images (objects, with hands and toothbrush activities with faces) were selected as neutral images (Table 1).

**Table 1.** The number and types of selected images (n=140) within each category and subdivisions.
- *Valence and Arousal Scales:* The valence and arousal rating scales of the Self-Assessment Manikin (SAM) were used to assess the emotional valence and arousal levels associated with each presented image. In the pilot study, participants rated valence and arousal on a 5-point Likert scale ranging from “1” to “5”. For the main study, a more detailed 9-point Likert scale was employed. On the valence scale, a minimum value of 1 was represented by a frowning, unhappy figure, indicating extreme unpleasantness, while the maximum value (5 or 9) was represented by a smiling, happy figure, representing extreme pleasantness (Bradley & Lang, 1994). The minimum value (1) on the arousal scale was accompanied by a relaxed and sleepy figure, indicating a feeling of calmness, while the maximum value (5 or 9) was accompanied by an excited, wide-eyed figure, corresponding to feeling very excited and aroused (Bradley & Lang, 1994). Participants were instructed to rate their responses after being presented with the stimulus, providing valuable insight into the emotional responses elicited by the images.
- *Craving:* In this study, we used two measures of craving, including the Visual Analog Scale (VAS) and the Desires for Drug Questionnaire (DDQ) (Franken et al., 2002). The VAS was used to visually measure the immediate desire for cannabis by participants in response to each image presented in both the pilot and online behavioral studies. A 0–100 mm VAS was used to determine the intensity of cue-induced craving, where 0 indicated “no craving” and 100 indicated “extreme craving”. Inside the scanner, participants were asked to rate their craving on a 4-point Likert scale, where 1 represented “not at all” and 4 represented “extremely” after each stimulus presentation. The DDQ is a self-report questionnaire comprising three subscales: desire and intention (7 questions), negative reinforcement (4 questions), and control (2 questions). This questionnaire has been validated for Iranian heroin users (Hassani-Abharian et al., 2016) and is widely used for different types of substances. Each question is rated on a 7-level Likert scale, where a score of 1 represents “completely disagree” and a score of 7 represents “completely agree “. In this study, we used the DDQ before and after the fMRI scanning session.
- *Oral Fluid Test sample:* Before each scanning session, a Six Panel multi-drug Saliva test kit (WONDFO biotech, USA) was used to screen the participants’ substance use. This test kit screened for the presence of amphetamines, methamphetamine, methadone, morphine, benzodiazepine, and cannabis. This screening ensured that participants were not poly-drug users, enhancing the reliability of the study results.
- *Cue reactivity fMRI paradigm:* A visual fMRI cannabis cue-reactivity task was designed to examine differences in activation for cannabis vs. non-cannabis neutral images (toothbrush-related images) (Karoly et al., 2019; Vollstädt-Klein et al., 2011). From the 140 images rated in the online behavioral study, 40 cannabis-related images (with the highest craving scores) and 40 neutral images (toothbrush-related images with the lowest craving scores) were selected for use in the DCR task. All images were of high resolution and scaled to similar dimensions to ensure a high-quality display in the MRI environment. After 24 seconds of resting-state, participants viewed images presented for 6 seconds in a random order, followed by a 4-second fixation period. Subsequently, participants rated their craving for the presented image on a scale from 1 to 4 (1 = not at all to 4 = extremely) using an MRI-compatible response box placed under both hands (duration was 6 seconds). Each trial lasted 16 seconds, and the total duration of the fMRI task was 664 seconds. Functional MRI images were collected using a Siemens MAGNETOM Prisma 3.0T scanner at the National Brain Mapping Laboratory. At first, we acquired a T1-weighted magnetization prepared rapid acquisition gradient echo (MPRAGE) sequence of 4 min 12 sec (160 sagittal slices; repetition time (TR) = 1800 ms; echo time (TE) = 3.53 ms; inversion time (TI) = 1100 ms; flip angle (FA) = 7°; slice thickness = 1.0 mm; field of view (FOV) = 256 mm; voxel size = 1 × 1 × 1 mm). The T2*-weighted gradient echo planar (EPI) sequence was acquired with 43 transversal slices oriented parallel to the AC–PC line (TR = 2000 ms; TE = 50 ms; FA = 90°; slice thickness 3.0 mm; FOV = 192 mm; voxel size = 3 × 3 × 3 mm) (TR = 2000 ms; TE = 50 ms; FA = 90°; slice thickness = 3.0 mm; FOV = 192 mm; voxel size = 3 × 3 × 3 mm).

| Categories (n) | Subdivisions (n) |
| --- | --- |
| Cannabis alone (20) | Cannabis flower (10) |
|  | Cannabis powder (10) |
| Cannabis-related paraphernalia objects (30) | Blunt/Joint objects (10) |
|  | Pipe/Bowl objects (10) |
|  | Bong objects (10) |
| Cannabis-related paraphernalia with hands (30) | Blunt/Joint with hand (10) |
|  | Pipe/Bowl with hand (10) |
|  | Bong with hand (10) |
| Cannabis-related paraphernalia activities with faces (30) | Blunt/Joint activities with faces (10) |
|  | Pipe/Bowl activities with faces (10) |
|  | Bong activities with faces (10) |
| Neutral (toothbrush) (30) | Toothbrush objects (10) |
|  | Toothbrush with hands (10) |
|  | Toothbrush activities with faces (10) |

### 2.6. Data Analysis

To ensure that each image elicited at least moderate craving, a one-sample t-test was used to compare each image’s mean craving rating to 50, which represents the “moderate” point on the craving scale. Similarly, valence and arousal ratings were compared to 5, representing the “moderate” point on the valence and arousal scale, respectively. Behavioral data were analyzed using SPSS (Statistical Package of the Social Sciences, Version 15.0.0, SPSS, Inc., Chicago, Illinois).

The AFNI software package was used to preprocess the functional MRI data (National Institute of Mental Health, Bethesda, MD; https://afni.nimh.nih.gov/). The first three functional scans were discarded to ensure steady-state magnetization. The preprocessing pipeline includes despiking, slice-time correction, realignment, co-registration, spatial normalization to the Montreal Neurological Institute (MNI) standardized space, and spatial smoothing with a 4-mm full-width at half-maximum Gaussian kernel. Times of repetition (TRs) with motion above 3 mm were censored.

The preprocessed fMRI data were analyzed using a general linear model (GLM) created by modeling onset times for the cannabis conditions and for the neutral conditions with a 6-second boxcar function, convolved with a standard hemodynamic response function (HRF) to generate two regressors of interest. Six motion correction parameters from each subject were included in the first-level model as nuisance regressors. The differential contrasts directly comparing the cannabis with the neutral conditions were included for each subject in second-level mixed-effects models developed using AFNI’s 3dMEMA. Based on Monte-Carlo simulations conducted in AFNI’s 3dClustSim, all group-level results were cluster-level corrected for multiple comparisons (P < 0.05, cluster size > 60).

### 2.7. Group Factor Analysis

We used group factor analysis (GFA) to investigate potential relationships between groups of variables with a sparsity constraint. GFA employs a sparse Bayesian estimation to find latent variables that either reflect a robust relationship between groups or explain away group-specific variation. Three variable groups were defined: (1) neural measures; (2) behavioral measures; and (3) demographic measures. For neural measures, cannabis minus neutral contrasts from 38 regions of interest (ROIs) (orbitofrontal cortex (47o_left, 47o_right, A11l_left, A11l_right), cingulate gyrus (A23c_left, A23c_right, A32p_left, A32p_right, A32sg_left, A32sg_right), precuneus (A31_left, A31_right, A5m_left, A5m_right, A7m_left, A7m_right, dmPOS_left, dmPOS_right), hippocampus (cHipp_left, cHipp_right, rHipp_left, rHipp_right), amygdala (lAmyg_left, lAmyg_right, mAmyg_left, mAmyg_right), basal ganglia (NAc_left, NAc_right, vCa_left, vCa_right, vmPu_left, vmPu_right, dlPu_left, dlPu_right), and insula (vIa_left, vIa_right, vIg_left, vIg_right), based on the results of meta-analysis were included as neural GFA group (Sehl et al., 2021).

The behavioral group consisted of 12 measures, including DDQ subscales (*Desire and intention, Negative reinforcement, Deficit of control*) both before and after scanning, as well as craving/thought/need self-reports before and after scanning. The demographic group comprised four measures: Age, Education, Cannabis use frequency, and Beck score. Therefore, the model included 38 ROI brain activation measures, 12 behavioral measures, and 4 demographic measures. The variables were z-normalized to have a zero mean and unit variance in order to provide a form appropriate for GFA. The GFA estimation process was repeated ten times to ensure the consistency of robust latent factors across the sample chains, minimizing the risk of identifying spurious latent factors.

We assessed potential bivariate relationships between neural and behavioral variables using Pearson’s correlation tests. This served as a less reliable but complementary test for neuro-behavioral associations. Pearson’s correlations and group factor analysis were conducted in statistical software R version 4.0.5. The GFA was conducted using the “gfa” function from the GFA package in R programming language.

## 3. Result

### 3.1. Pilot Study

#### 3.1.1. Demographic and Cannabis Use Descriptive Data

In the pilot phase, 10 participants completed the single face-to-face session, with a mean age of 19.71 years (SD = 6.8). Of these, 3 participants were female and 7 were male at the Bachelor’s (n = 7) and Master’s (n = 3) degree levels. The mean age of onset for cannabis use was 18.5 (SD = 2.9) years, with an average of 4.22 (SD = 3.3) years of regular cannabis use.

#### 3.1.2. Image Rating

Table 2 shows the mean values (and standard deviations) of valence, arousal, and craving for each category, including specific methods of use, and each subdivision, representing specific drug-related actions. It should be noted that images indicating cannabis vapor were not rated by participants and were indicated as “*Not Applicable*” since they were an unfamiliar and uncommon method of using cannabis among Iranian users. According to the pilot study, we selected the subcategories that elicited higher subjective craving scores from the participants.

**Table 2.** Mean values of valence, arousal, and craving scores for cannabis-related images in the pilot study for all 356 images.

| Subdivisions (n) | Categories (specific use method)<br>(n) | Craving (1-5)<br>Mean (SD) | Arousal (1-5)<br>Mean (SD) | Valence (1-5)<br>Mean (SD) |
| --- | --- | --- | --- | --- |
| Cannabis-related<br>paraphernalia objects<br>(127) | Cannabis flower (32) | 4.15 (0.9) | 4.01 (1.23) | 3.51 (1.01) |
|  | Pipe/Bowl (27) | 1.99 (1.02) | 1.99 (0.92) | 3.21 (0.98) |
|  | Bong (24) | 1.15 (0.32) | 2.1 (1.23) | 2.5 (0.45) |
|  | Blunt/Joint (24) | 2.7(1.3) | 3.739 (1.02) | 3.49 (0.91) |
|  | Vapor (20) | NA | NA | NA |
| Cannabis-related<br>paraphernalia with hands<br>(82) | Pipe/Bowl (20) | 2.04 (0.94) | 3.01 (1.41) | 3.31 (0.77) |
|  | Bong (21) | 1.33 (0.73) | 2.29 (1.14) | 2.11 (0.89) |
|  | Blunt/Joint (21) | 2.96(1.16) | 3.32 (0.89) | 3.24 (1.11) |
|  | Vapor (20) | NA | NA | NA |
| Cannabis-related<br>paraphernalia activities with<br>faces<br>(87) | Pipe/Bowl (21) | 2.24 (1.01) | 2.11 (1.48) | 2.5 (1.03) |
|  | Bong (22) | 1.49 (0.58) | 2.31 (0.98) | 1.3 (1.21) |
|  | Blunt/Joint (24) | 3.04 (1.04) | 3.9 (1.11) | 3.59 (0.42) |
|  | Vapor (20) | NA | NA | NA |

NA= Not Applicable; SD= Standard Deviation.

### 3.2. Behavioral Study (Cue Validation)

#### 3.2.1. Demographic and Cannabis Use Characteristics Data

The demographics and cannabis use characteristics of participants in the main study are summarized in Table 3.

**Table 3.**
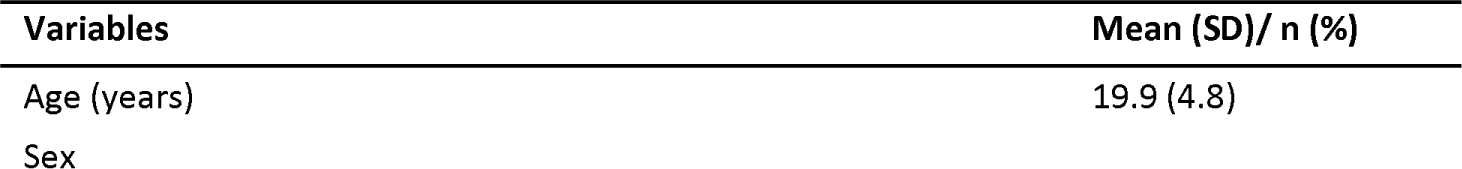
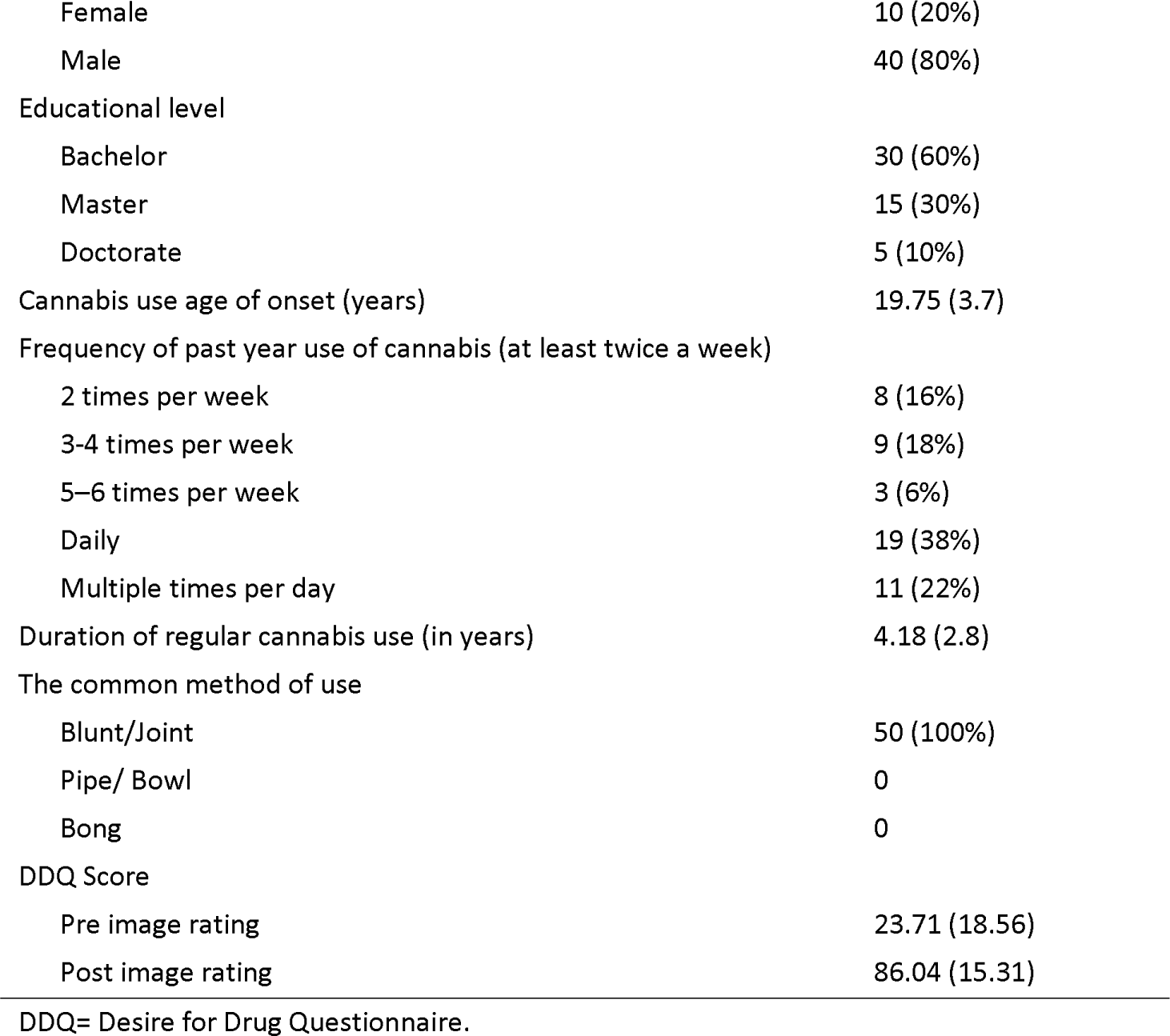
Participants’ demographics and substance use characteristics in the main study (n = 50).

#### 3.2.2. Images Rating

Mean values (SD) of ratings for each subdivision in terms of craving, valence, and arousal are presented in Tables 4 and 5, respectively, for cannabis-related (n = 110) and control (n = 30) images.

**Table 4.** Craving, arousal, valence values for cannabis-related images within each category and each subdivision.

| Categories and subdivisions (n) | Craving (0-100)<br>Mean (SD) | Arousal (1-9)<br>Mean (SD) | Valence (1-9)<br>Mean (SD) |
| --- | --- | --- | --- |
| Cannabis-related images (Total=110) | 53.9 (16.72) | 4.49 (13.2) | 5.43 (3.2) |
| Cannabis alone (20) | 59.12 (5.11) | 5.2 (0.4) | 5.67 (1.1) |
| Cannabis flower (10) | 60.88 (6.25) | 5.31 (0.29) | 6.49 (0.20) |
| Cannabis powder (10) | 52.30 (10.55) | 5.11 (0.37) | 4.9 (1.21) |
| Cannabis-related paraphernalia objects |  |  |  |
| Blunt/Joint objects (10) | 56.29 (4.22) | 5.12 (0.57) | 6.12 (0.31) |
| Pipe/Bowl objects (10) | 38.16 (7.01) | 4.15 (0.4) | 5.47 (0.4) |
| Bong objects (10) | 33.6 (8.7) | 3.2 (0.4) | 5.01 (0.40) |
| Cannabis-related paraphernalia with hands |  |  |  |
| Blunt/Joint with hands (10) | 52.55 (7.01) | 5.41 (0.7) | 5.83 (0.32) |
| Pipe/Bowl with hands (10) | 34.9 (7.51) | 3.9 (0.38) | 5.31 (0.53) |
| Bong with hands (10) | 35.45 (1.54) | 3.89 (0.25) | 5.21 (0.19) |
| Cannabis-related paraphernalia activities with faces |  |  |  |
| Blunt/Joint activities with faces (10) | 51.58 (6.99) | 5.54 (0.44) | 5.32 (0.38) |
| Pipe/Bowl activities with faces (10) | 47.2 (7.1) | 3.9 (0.41) | 5.06 (0.39) |
| Bong activities with faces (10) | 39.9 (3.6) | 3.89 (0.4) | 5.14 (0.5) |

**Table 5.** Craving, arousal, valence values for neutral (toothbrush) images.

| Neutral Images (n) | Craving (1-100) | Arousal (1-9) | Valence (1-9) |
| --- | --- | --- | --- |
|  | Mean (SD) | Mean (SD) | Mean (SD) |
| Toothbrush images | 15.45 (2.9) | 1.45 (0.4) | 5.06 (1.19) |
| Toothbrush-related paraphernalia objects (10) | 15.04 (3.4) | 1.5 (0.5) | 4.99 (1.4) |
| Toothbrush-related paraphernalia with hands (10) | 16.15 (2.1) | 1.7 (0.14) | 5.1 (1.2) |
| Toothbrush-related paraphernalia activities with faces (10) | 15.18 (2.96) | 1.22 (0.3) | 5.1 (1.14) |

One-sample t-tests were used to compare each image’s mean craving rating to 50 (moderate) in each category and subdivision(Macatee et al., 2021). The results showed that the subdivisions of Cannabis powder (t = 3.79 and *p* = 0.004), Cannabis flower (t = 5.67 and *p*<0.001), Blunt/Joint objects (t = 4.99 and *p* = 0.001), Blunt/Joint with hands (t = 4.49 and *p* = 0.002), and Blunt/Joint activities with faces (t = 3.47 and *p* = 0.007) had significant craving scores compared to the moderate point, inducing at least moderately intense craving in participants. The results are presented in Table 6.

**Table 6.** The comparison of each individual image’s mean craving rating with 50 (moderate) and mean the difference.

| Categories and subdivisions (n) | Craving (moderate=50) |  |  |
| --- | --- | --- | --- |
|  | t | p-value | Mean difference |
| Cannabis-related images (110) | 2.2 | 0.2 | -1.2 |
| Neutral (toothbrush) (30) | -46.18 | < 0.001 | -33.7 |
| Cannabis alone (20) | 6.66 | < 0.001 | 10.3 |
| Blunt/Joint (30) | 7.36 | < 0.001 | 6.3 |
| Pipe /Bowl (30) | -6.45 | < 0.001 | -6.7 |
| Bong (30) | -9.91 | < 0.001 | -8.61 |
| Cannabis alone |  |  |  |
| Cannabis powder (10) | 3.79 | $p = 0.004$ | 9.8 |
| Cannabis flower (10) | 5.67 | < 0.001 | 12.8 |
| Cannabis-related paraphernalia objects |  |  |  |
| Blunt/Joint objects (10) | 4.99 | $p = 0.001$ | 7.9 |
| Pipe /Bowl objects (10) | -5.88 | < 0.001 | -18.9 |
| Bong objects (10) | -3.83 | $p = 0.004$ | -11.55 |
| Toothbrush objects (10) | -21.01 | $< 0.001$ | -34.6 |
| Cannabis-related paraphernalia with hands |  |  |  |
| Blunt/Joint with hands (10) | 4.49 | $p = 0.002$ | 5.88 |
| Pipe /Bowl with hands (10) | -11.61 | $< 0.001$ | -24.1 |
| Bong with hands (10) | -16.42 | $< 0.001$ | -13.2 |
| Toothbrush with hands (10) | -32.99 | $< 0.001$ | -35.3 |
| Cannabis-related paraphernalia activities with faces |  |  |  |
| Blunt/Joint activities with faces (10) | 3.47 | $p = 0.007$ | 5.91 |
| Pipe /Bowl activities with faces (10) | -0.84 | 0.42 | -14.31 |
| Bong activities with faces (10) | -5.09 | 0.03 | -11.4 |
| Toothbrush activities with faces (10) | -32.18 | $< 0.001$ | -34 |

Similarly, one-sample t-tests were used to compare each individual image’s mean arousal score to 5 (moderate) in each category and subdivision (Macatee et al., 2021). The results showed that the subdivisions of Cannabis powder (t = 2.62 and *p* = 0.027), Cannabis flower (t = 4.72 and *p* = 0.02), Blunt/Joint objects (t = 5.98 and *p* < 0.001), Blunt/Joint with hands (t = 2.4 and *p* = 0.026), and Blunt/Joint activities with faces (t = 3.47 and *p* = 0.006) had a mean arousal rating significantly higher than 5, indicating that all images in these categories elicited at least moderately intense arousal. The results are presented in Table 7.

**Table 7.** The comparison of each individual image’s mean arousal rating with 5 (moderate) and the mean the difference.

| Categories and Subdivisions (n) | Arousal (moderate= 5) |  |  |
| --- | --- | --- | --- |
|  | t | p-value | Mean Difference |
| Cannabis related images (110) | -7.6 | 0.003 | -0.2 |
| Neutral (toothbrush) (30) | -44.21 | $< 0.001$ | -3.26 |
| Cannabis alone (20) | 4.23 | $< 0.001$ | 0.45 |
| Blunt/Joint (30) | 6.77 | $< 0.001$ | 0.75 |
| Pipe /Bowl (30) | -12.23 | $< 0.001$ | -1.2 |
| Bong (30) | -14.21 | $< 0.001$ | -1.1 |
| Cannabis alone |  |  |  |
| Cannabis powder (10) | 2.62 | 0.027 | 0.48 |
| Cannabis flower (10) | 4.72 | 0.002 | 0.42 |
| Cannabis-related paraphernalia objects |  |  |  |
| Blunt/Joint objects (10) | 5.98 | $< 0.001$ | 1.16 |
| Pipe /Bowl objects (10) | -5.4 | $< 0.001$ | -0.75 |
| Bong objects (10) | -8.19 | $< 0.001$ | -1.19 |
| Toothbrush objects (10) | -19.32 | $< 0.001$ | -3.4 |
| Cannabis-related paraphernalia with hands |  |  |  |
| Blunt / Joint with hands (10) | 2.46 | 0.026 | 0.58 |
| Pipe / Bowl with hands (10) | -8.41 | < 0.001 | -1.10 |
| Bong with hands (10) | -12.31 | < 0.001 | -1.11 |
| Toothbrush with hands (10) | -63.43 | < 0.001 | -3.07 |
| Cannabis-related paraphernalia activities with faces |  |  |  |
| Blunt/Joint activities with faces (10) | 3.47 | 0.006 | 0.51 |
| Pipe /Bowl activities with faces (10) | -8.4 | < 0.001 | -1.18 |
| Bong with activities faces (10) | -6.11 | < 0.001 | -1.03 |
| Toothbrush activities with faces (10) | -30.43 | < 0.001 | -3.12 |

Furthermore, one-sample t-tests were used to compare each individual image’s mean valence score to 5 (moderate) in each category (Macatee et al., 2021). The results showed that the category of cannabis-related images (t = 4.44 and *p* < 0.001) and the subdivisions of Cannabis powder (t = 3.31 and *p* = 0.009), Cannabis flower (t = 23.61 and *p* < 0.001), Blunt/Joint objects (t = 12.16 and *p* < 0.001), Pipe/Bowl objects (t = 3.61 and *p* = 0.005), Blunt/Joint with hands (t = 7.91 and *p* < 0.001), Bong with hands (t = 3.46 and *p* = 0.007), and Blunt/Joint activities with faces (t = 4.33 and *p* = 0.002) had a mean valence score significantly higher than 5, indicating that all images in these categories elicited at least moderately intense valence. The results are presented in Table 8.

**Table 8.**
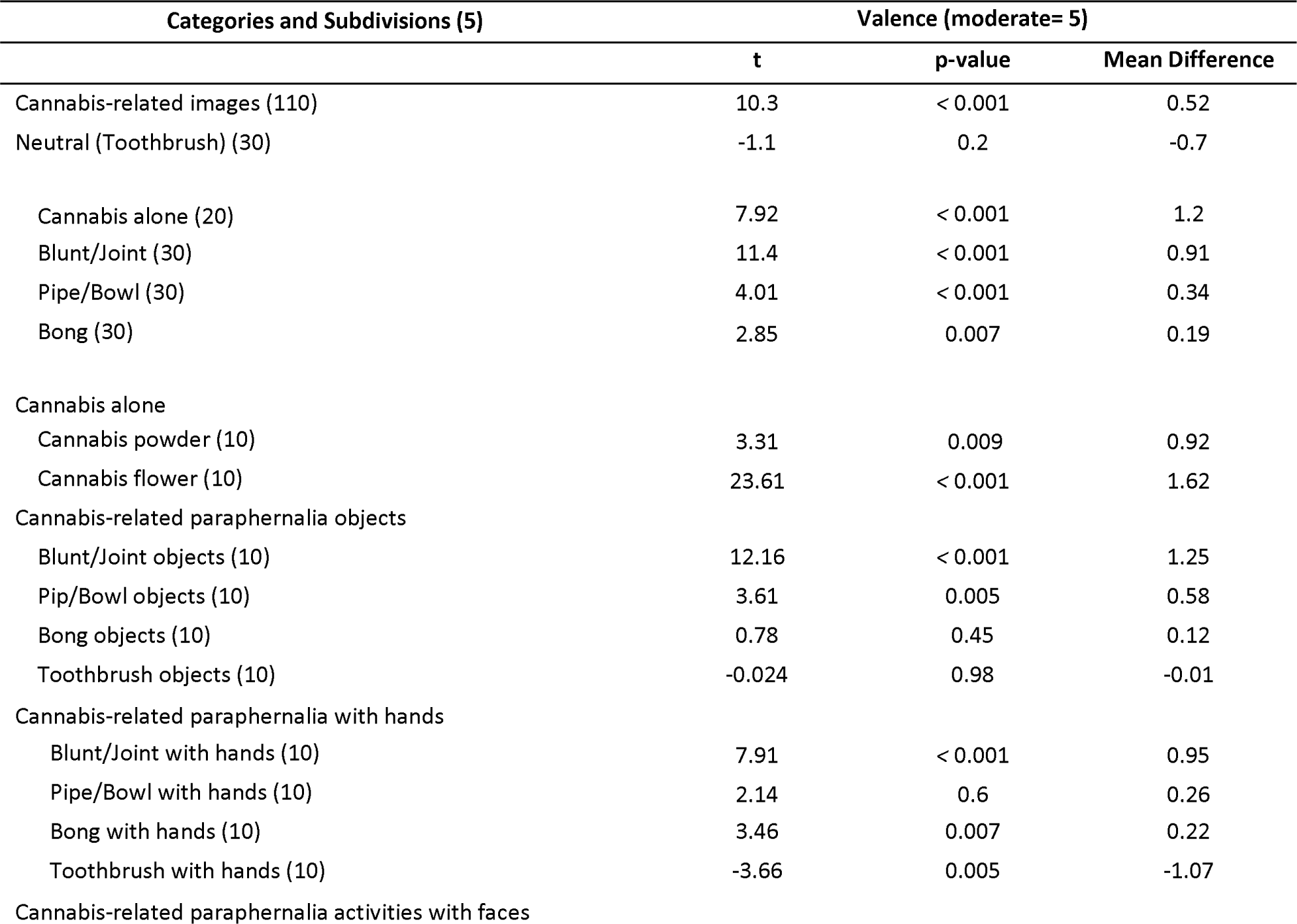
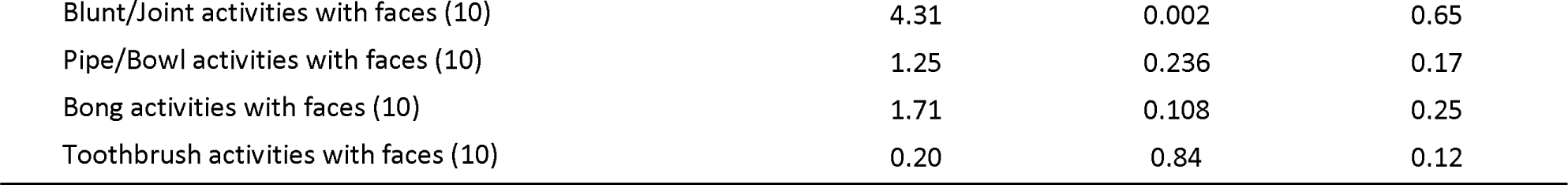
The comparison of each individual image’s mean valence rating to 5 (moderate) and the mean the difference.

| Categories and Subdivisions (5) | Valence (moderate= 5) |  |  |
| --- | --- | --- | --- |
|  | t | p-value | Mean Difference |
| Cannabis-related images (110) | 10.3 | < 0.001 | 0.52 |
| Neutral (Toothbrush) (30) | -1.1 | 0.2 | -0.7 |
| Cannabis alone (20) | 7.92 | < 0.001 | 1.2 |
| Blunt/Joint (30) | 11.4 | < 0.001 | 0.91 |
| Pipe/Bowl (30) | 4.01 | < 0.001 | 0.34 |
| Bong (30) | 2.85 | 0.007 | 0.19 |
| Cannabis alone |  |  |  |
| Cannabis powder (10) | 3.31 | 0.009 | 0.92 |
| Cannabis flower (10) | 23.61 | < 0.001 | 1.62 |
| Cannabis-related paraphernalia objects |  |  |  |
| Blunt/Joint objects (10) | 12.16 | < 0.001 | 1.25 |
| Pip/Bowl objects (10) | 3.61 | 0.005 | 0.58 |
| Bong objects (10) | 0.78 | 0.45 | 0.12 |
| Toothbrush objects (10) | -0.024 | 0.98 | -0.01 |
| Cannabis-related paraphernalia with hands |  |  |  |
| Blunt/Joint with hands (10) | 7.91 | < 0.001 | 0.95 |
| Pipe/Bowl with hands (10) | 2.14 | 0.6 | 0.26 |
| Bong with hands (10) | 3.46 | 0.007 | 0.22 |
| Toothbrush with hands (10) | -3.66 | 0.005 | -1.07 |
| Cannabis-related paraphernalia activities with faces |  |  |  |
| Blunt/Joint activities with faces (10) | 4.31 | 0.002 | 0.65 |
| Pipe/Bowl activities with faces (10) | 1.25 | 0.236 | 0.17 |
| Bong activities with faces (10) | 1.71 | 0.108 | 0.25 |
| Toothbrush activities with faces (10) | 0.20 | 0.84 | 0.12 |

Additionally, independent samples t-test was used to compare craving, arousal, and valence between cannabis-related images and neutral images. The results showed significant differences between all categories and subdivisions with neutral images. Specifically, the category of cannabis-related paraphernalia with hands was the only one that showed significant differences with neutral (t = 6.99 and *p* < 0.001). The results are presented in Table 9.

**Table 9.** The comparison of craving, arousal, and valence between cannabis-related images and neutral (toothbrush) images.

| Categories and Subdivisions (n) | Craving (1-50) |  | Arousal (1-5) |  | Valence (1-5) |  |
| --- | --- | --- | --- | --- | --- | --- |
|  | t | p-value | t | p-value | t | p-value |
| Cannabis related images (110) vs. neutral (toothbrush) (30) | 6.56 | < 0.001 | 17.46 | < 0.001 | 2.67 | 0.08 |
| Cannabis (20) vs. neutral (toothbrush) (30) | 20.9 | < 0.001 | 19.35 | < 0.001 | 3.27 | < 0.001 |
| Blunt/Joint (30) vs. neutral (toothbrush) (30) | 26.19 | < 0.001 | 23.25 | < 0.001 | 4.22 | < 0.001 |
| Pipe/Bowl (30) vs. neutral (toothbrush) (30) | 18.77 | < 0.001 | 19.81 | < 0.001 | 2.21 | 0.32 |
| Bong (30) vs. neutral (toothbrush) (30) | 20.40 | < 0.001 | 19.86 | < 0.001 | 1.71 | 0.8 |
| Cannabis-related paraphernalia objects (30) vs. neutral (toothbrush) objects (10) | 9.53 | < 0.001 | 8.28 | < 0.001 | 1.81 | 0.06 |
| Blunt/Joint objects (10) vs. neutral (toothbrush) objects (10) | 18.61 | < 0.001 | 9.9 | < 0.001 | 0.5 | 0.62 |
| Pipe/Bowl objects (10) vs. neutral (toothbrush) objects (10) | 7.91 | < 0.001 | 10.88 | < 0.001 | 0.53 | 0.59 |
| Bong objects (10) vs. neutral (toothbrush) objects (10) | 9.37 | < 0.001 | 9.71 | < 0.001 | 0.22 | 0.82 |
| Cannabis-related paraphernalia with hands (30) vs. neutral (toothbrush) with hands (10) | 11.18 | < 0.001 | 8.28 | < 0.001 | 6.99 | < 0.001 |
| Blunt/Joint with hands (10) vs. neutral (toothbrush) with hands (10) | 24.08 | < 0.001 | 2.21 | $p = 0.04$ | 0.33 | 0.74 |
| Pipe/Bowl with hands (10) vs. neutral (toothbrush) with hands (10) | 4.69 | < 0.001 | 6.36 | < 0.001 | 4.64 | 0.001 |
| Bong with hands (10) vs. neutral (toothbrush) with hands (10) | 19.47 | < 0.001 | 12.89 | < 0.001 | -3.32 | 0.002 |
| Cannabis-related paraphernalia activities with faces (30) vs. neutral with faces (10) | 12.09 | < 0.001 | 8.97 | < 0.001 | 0.66 | 0.53 |
| Blunt/Joint activities with faces (10) vs. neutral (toothbrush) activities with faces (10) | 19.42 | < 0.001 | 6.46 | < 0.001 | 0.55 | 0.4 |
| Pipe/Bowl activities with faces (10) vs. neutral (toothbrush) activities with faces (10) | 10.31 | < 0.001 | 13.22 | < 0.001 | 0.72 | 0.47 |
| Bong activities with faces (10) vs. neutral (toothbrush) activities with faces (10) | 16.69 | < 0.001 | 2.91 | $p = 0.04$ | -2.05 | 0.54 |

Correlation analysis showed no significant correlations between craving and reaction time for cannabis (R = 0.09, p = 0.3; Pearson’s correlation) and neutral (R = -0.36, p = 0.051; Pearson’s correlation) cues (Figure 2A).

**Figure 2.**
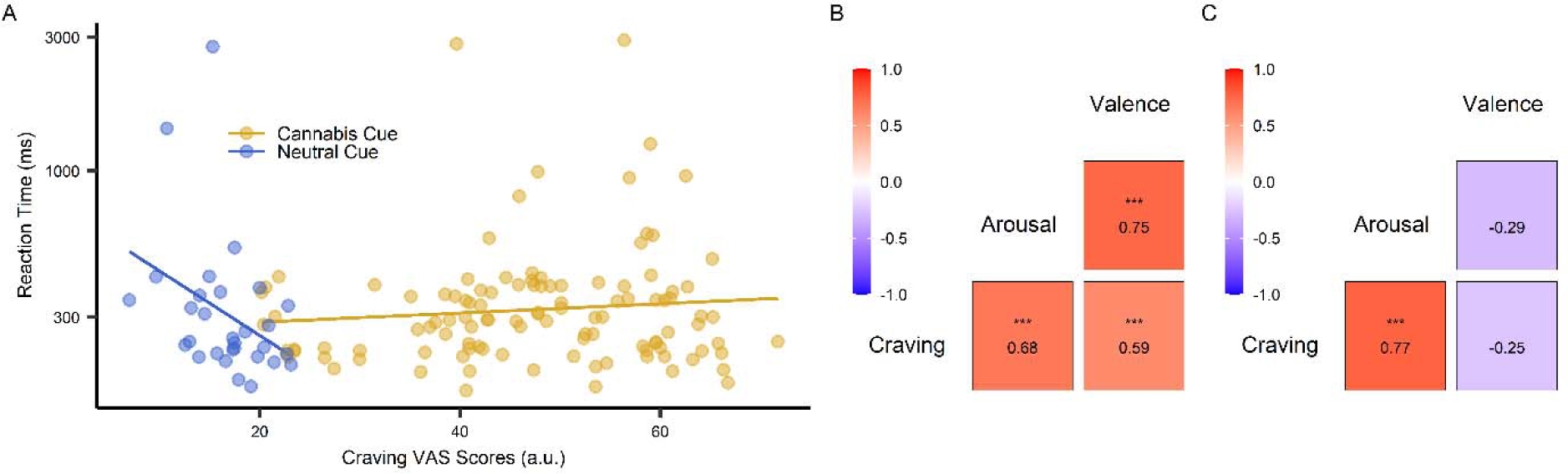
Relations of behavioral responses to pictorial cannabis and neutral cues. **(A)** Correlation between reaction time and craving scores. The scatterplot represents the relationship between reaction time and craving for cannabis (R = 0.09; p = 0.3; Pearson’s correlation) and neutral (R = -0.36; p = 0.051; Pearson’s correlation) cues. Each point presents data from the participants’ average responses to each individual picture. **(B,C)** The corresponding correlation matrices between craving, valence, arousal for cannabis (B) and neutral cues (C).

Furthermore, we tested for bivariate correlations between psychological variables including craving, arousal, and valence. These tests revealed significant positive correlations between arousal and craving scores for cannabis (r = 0.68, p < 0.001; Spearman’s correlation) (Figure 2B) and neutral (r = 0.77, p < 0.001; Spearman’s correlation) cues (Figure 2C). Other significant correlations between valence and craving scores (r = 0.59, p < 0.001; Spearman’s correlation) and between valence and arousal scores (r = 0.75, p < 0.001; Spearman’s correlation) within cannabis cues (Figure 3B). Moreover, there were no significant correlations between valence and craving scores (r = -0.25, p = 0.18; Spearman’s correlation) and between valence and arousal scores (r = -0.29, p = 0.11; Spearman’s correlation) within neutral cues (Figure 2C). The distribution of craving in each category of pictures is shown in Figure 3.

**Figure 3.**
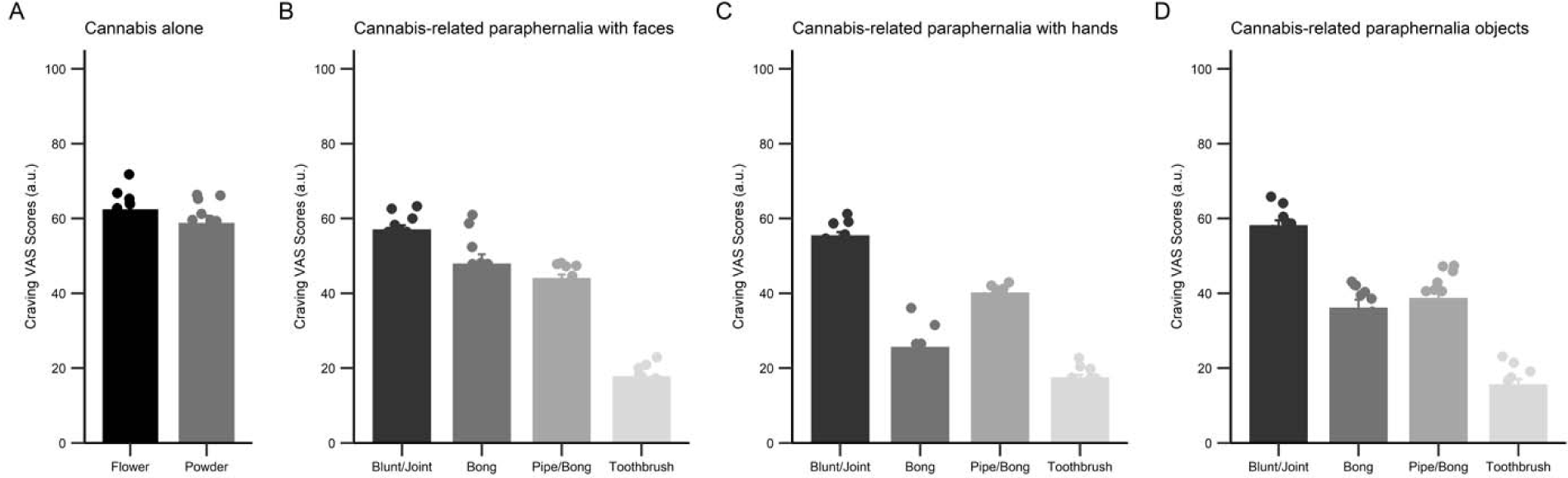
Distribution of craving scores in four categories of the pictures. Representative bar charts showing craving scores in four categories of the pictures (A) cannabis alone; (B) Cannabis-related paraphernalia with faces; (C) Cannabis-related paraphernalia with hands; and (D) Cannabis-related paraphernalia objects. Data in bar charts are represented as mean ± SEM.

### 3.3. Neural Study (Cannabis Cue Reactivity)

#### 3.3.1. Demographic and Cannabis Use Descriptive in the Main Study

Thirteen participants were excluded from fMRI analyses due to positive COVID test result. In addition, four participants were excluded due to excessive motion (>3 mm), and two participants could not complete the fMRI task. The remaining sample consisted of 31 cannabis users. The demographics and cannabis use characteristics of participants in the fMRI study are presented in Table 10.

**Table 10.** Participants’ demographics and substance use characteristics in the fMRI study (n = 31). Values are reported as mean (SD)/frequency (%).

| Variables | Values |
| --- | --- |
| Age (years) | 20.18 (3.34) |
| Sex |  |
| Female | 4 (12.9%) |
| Male | 27 (87.09%) |
| Educational level |  |
| Bachelor | 20 (64.51%) |
| Master | 10 (32.25%) |
| Doctorate | 1 (3.25%) |
| Cannabis use age of onset (years) | 29.09 (3.55) |
| Frequency of past year use of cannabis (at least twice a week in years) |  |
| 2 times per week | 8 (25.75%) |
| 3-4 times per week | 7 (22.58%) |
| 5-6 times per week | 5 (16.12%) |
| Daily | 6 (19.35%) |
| Multiple times per day | 5 (16.19%) |
| Duration of regular cannabis use (in years) | 4.91 (2.45) |
| The common method of use |  |
| Blunt/Joint | 31 (100%) |
| Pipe/ Bowl | 0 |
| Bong | 0 |

#### 3.3.2. Craving

To test whether cue exposure increased participants’ craving level, a paired sample t-test was used. Our results indicated that craving after the cue exposure task significantly increased craving levels (*p* < 0.001, t= 7.61).

#### 3.3.3. fMRI Analysis

To examine how cannabis cue-reactivity influenced the brain’s circuitry, we analyzed BOLD activity measured during the cannabis cue-reactivity task at the whole-brain level using a GLM analysis (see Figure 4 and Table 11).

**Figure 4.**
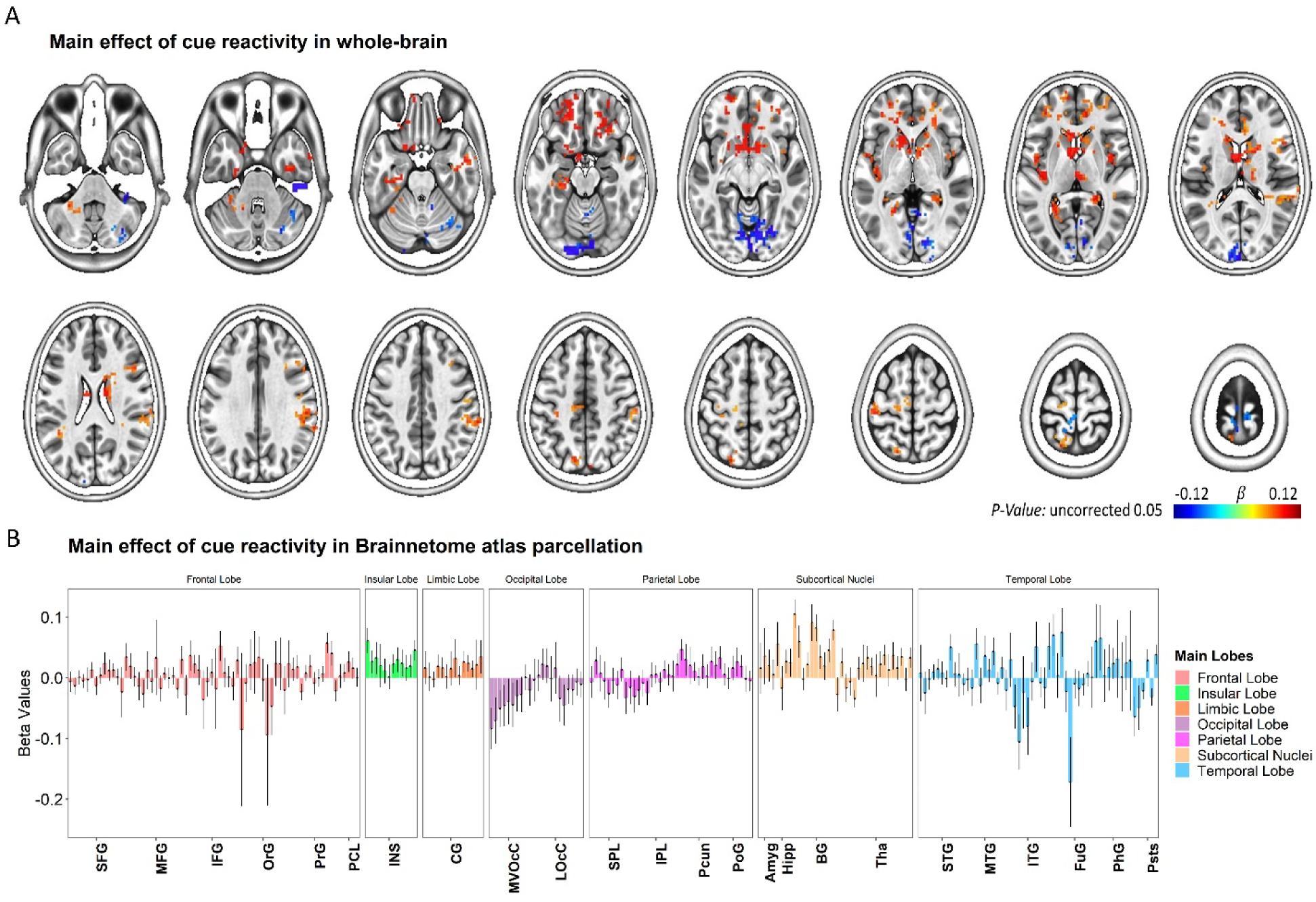
Whole-brain response to the task-based fMRI in contrasts of Cannabis > Neutral. **(A)** Brain activation maps and **(B)** changes in brain activation in Brainnetome (BNA) regions. Data in bar charts are represented as mean ± s.e.m. SFG, superior frontal gyrus; MFG, middle frontal gyrus; IFG: inferior frontal gyrus; OrG: orbital gyrus; PrG, precentral gyrus; PCL, paracentral lobule; STG, superior temporal gyrus; MTG, middle temporal gyrus; ITG, inferior temporal gyrus; FuG, fusiform gyrus; PhG, parahippocampal gyrus; pSTS, posterior superior temporal sulcus; SPL, superior parietal lobule; IPL, inferior parietal lobule; Pcun, precuneus; PoG, postcentral gyrus; INS, insular gyrus; CG, cingulate gyrus; MVOcC, medioventral occipital cortex; LOcC, lateral occipital cortex; Amyg, amygdala; Hipp, hippocampus; BG, basal ganglia; Tha, thalamus.

**Table 11.** Significant clusters for the main effects of cannabis cue reactivity in whole-brain analysis.

| Label | Side | Peak activation |  |  | Number of voxels | t-value |
| --- | --- | --- | --- | --- | --- | --- |
|  |  | x | y | z |  |  |
| Inferior frontal gyrus | L | 15 | -12 | -33 | 960 | 2.55 |
| Fusiform Gyrus | L | 21 | 93 | -21 | 407 | -5.74 |
| Parahippocampal Gyrus | R | -24 | 42 | 3 | 293 | 3.81 |
| Orbital Gyrus | L | 18 | -30 | -30 | 194 | 2.48 |
| Postcentral Gyrus | L | 45 | 27 | 63 | 140 | 2.78 |
| Insula | R | -42 | 0 | 6 | 127 | 4.32 |
| Precuneus | R | -6 | 81 | 48 | 86 | 2.24 |
| Superior Temporal Gyrus | L | 48 | 3 | 3 | 84 | 4.10 |
| Parahippocampal Gyrus | L | 24 | 54 | 6 | 77 | 2.96 |
| Cerebellar Tonsil | R | -45 | 39 | -57 | 75 | -2.72 |
| Declive | R | -33 | 66 | -27 | 75 | -3.76 |
| Culmen | L | 39 | 36 | -39 | 73 | 2.17 |
| Uncus | L | 24 | 12 | -36 | 67 | 2.99 |
| Medial Frontal Gyrus | R | -3 | 30 | 84 | 63 | -3.36 |
| Middle Temporal Gyrus | R | -63 | 0 | -33 | 62 | 3.90 |

*Note.* Whole-brain activations are clustered with a minimum cluster size of $k = 60$ , which corresponds to a cluster-level alpha of $p < 0.05$ using NN2 clustering. Abbreviation: L, left; R, right.

As expected, the main effect of cue-reactivity (contrast: cannabis > neutral) was significant in several clusters. These clusters included regions in the inferior/medial frontal gyrus, fusiform gyrus, parahippocampal gyrus, orbital gyrus, postcentral gyrus, insula, precuneus, superior/middle temporal gyrus, and cerebellar tonsil (see Figure 4A). In addition, we also reported the brain activation results across the 246 subregions in the human Brainnetome Atlas (see Figure 4B).

#### 3.3.4. Brain-Behavior Relationships

Two robust latent variables that collectively account for 15.12% of the variance across variable groups were found by employing group factor analysis (see Figure 5).

**Figure 5.**
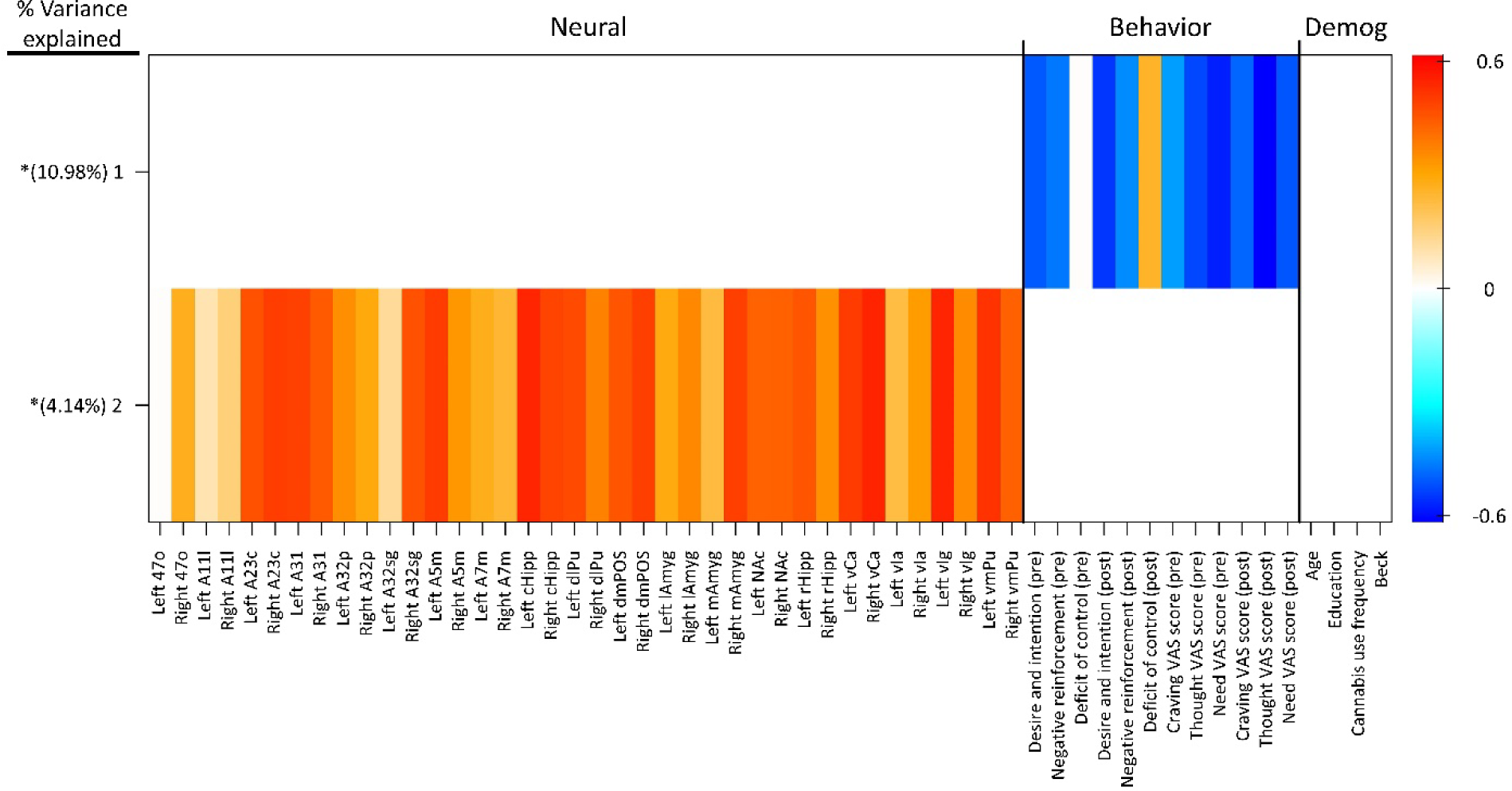
GFA robust factor loadings. Heatmap colors indicate the weight of each group variable loading. Robust group factors are sorted in descending order by mean % variance explained across all groups. Asterisks indicate group factors that contained at least one group variable loading whose 95% credible interval did not contain zero.

The mean-variance explained for the groups of behavioral and neural variables was 10.98 and 4.14%, respectively. There were no robust cross-unit latent variables identified between the neural group with behavioral and demographic groups. To put it another way, the GFA was unable to show any coherent relationship between the neural group with the behavioral and demographic groups in the latent variable space. In contrast, the significant bivariate relationships between neural and behavioral variables were found using the Pearson’s correlation tests as a less reliable, complementary test for neuro-behavioral associations (Figure 6A). These Pearson’s correlation tests included the individual BOLD signal changes (contrast: cannabis vs. neutral) in the regions of interest and behavioral parameters (defined as changes in total DDQ score or DDQ subscales (*Desire and intention, Negative reinforcement, Deficit of control*), post-fMRI – pre-fMRI). Here, the individual BOLD signal changes in the left A7m subregion were positively and significantly with overall DDQ (R = 0.38, P = 0.034; Figure 6A) and *Deficit of control* subscale (R = 0.4; P = 0.024; Figure 6K). The individual BOLD signal changes in the right vmPu subregion were correlated with overall DDQ (R = -0.44, P = 0.014; Figure 6E), *Desire and intention* subscale (R = -0.39, P = 0.029; Figure 6G), and *Negative reinforcement* (R = -0.36, P = 0.045; Figure 6J). The individual BOLD signal changes in the left dlPu subregion were negatively and significantly related to overall DDQ (R = -0.39; P = 0.029; Figure 6C) and *Negative reinforcement* (R = -0.44; P = 0.012; Figure 6H). The individual BOLD signal changes in the left mAmyg subregion were negatively and significantly related to overall DDQ (R = -0.46; P = 0.0098; Figure 6D) and *Negative reinforcement* (R = -0.39; P = 0.03; Figure 6I). There were other significant correlations between individual BOLD signal changes in the right cHipp subregion with overall DDQ (Figure 6B), in the right A32p subregion with *Desire and intention* subscale (Figure 6F), and in the right vIa subregion with *Deficit of control* subscale (Figure 6L).

**Figure 6.**
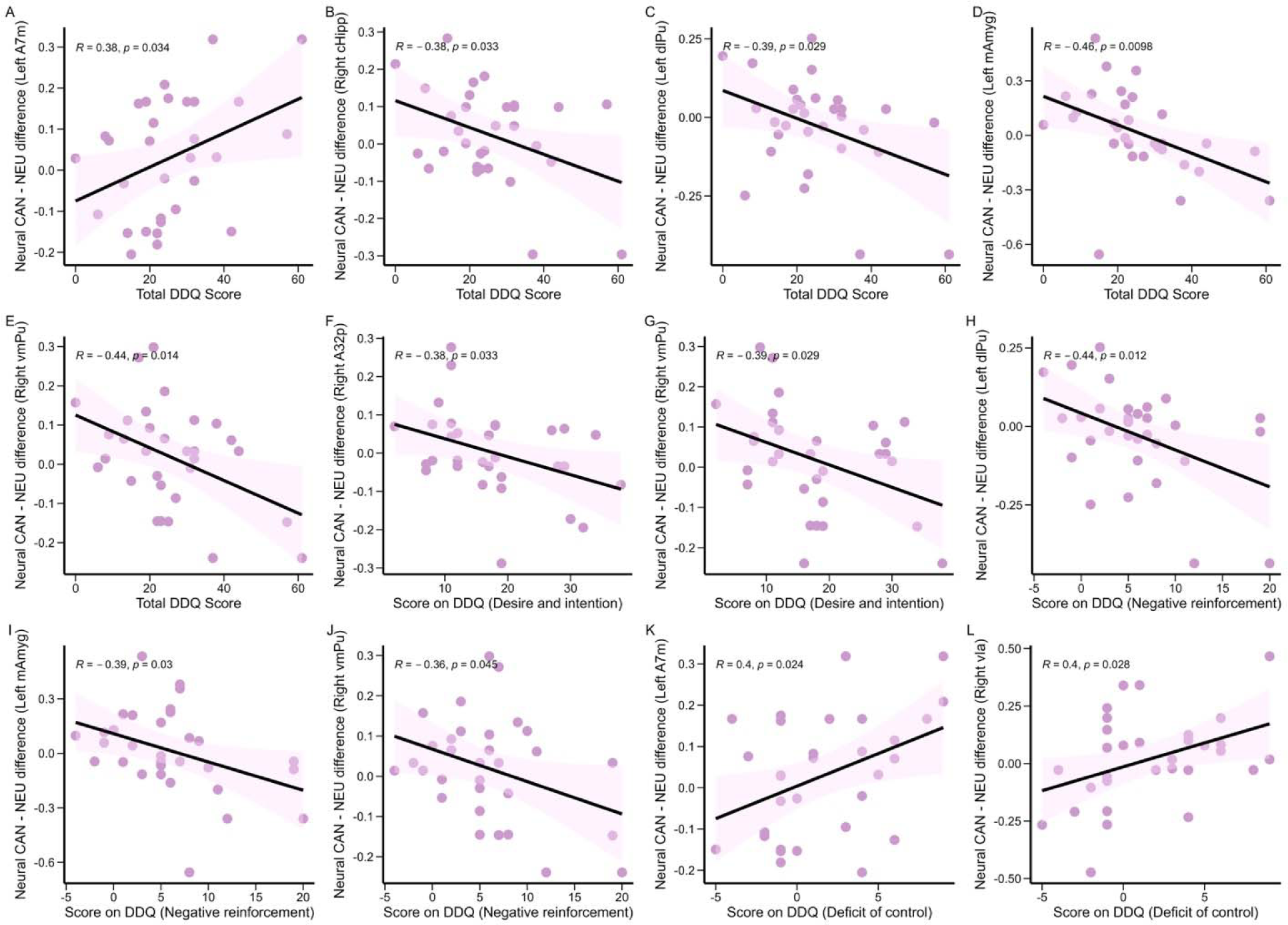
Correlations between neural and behavioral findings. Participants’ total scores on the DDQ correlated with individual BOLD signal changes in the left A7m (A), right cHipp (B), left dlPu (C), left mAmyg (D), and right vmPu (E). Participants’ scores on the Desire and intention subscale of the DDQ correlated with individual BOLD signal changes in the right A32p (F) and right vmPu (G). Participants’ scores on the Negative reinforcement subscale of the DDQ correlated with individual BOLD signal changes in the left dlPu (H), left mAmyg (I), and right vmPu (J). Participants’ scores on the Deficit of control subscale of the DDQ correlated with individual BOLD signal changes in the left A7m (K) and right vIa (L).

## 4. Discussion

Our study investigated cannabis cue reactivity in regular users using a combined behavioral and fMRI approach. We successfully identified and validated cannabis-related images capable of inducing craving and activating reward-related brain regions. These findings contribute to the understanding of neural mechanisms underlying cannabis cue reactivity and have potential implications for treatment development.

### 4.1. Specificity of Cue-Elicited Responses

Our study confirmed the potent nature of cannabis cues in triggering craving and motivational responses. Both behavioral data (significantly higher craving, arousal, and valence ratings) and fMRI data (increased activation in reward-related brain regions) provided converging evidence for cue-elicited reactivity. This aligns with previous research highlighting the ability of drug cues to elicit robust emotional and neurocognitive responses in individuals with SUDs (Ekhtiari et al., 2020; Sinha & Li, 2007; Volkow & Fowler, 2000). Notably, our study employed multiple measures to comprehensively assess cue reactivity, including subjective ratings, self-report questionnaires, and objective brain activity measures. This multimethod approach strengthens the confidence in our findings and offers a more comprehensive understanding of cue reactivity compared to studies relying solely on self-report measures.

### 4.2. Decoding Reward Circuitry Activation

The present study identified activation in reward-related brain regions during exposure to cannabis cues, including the frontal gyrus, insula, and hippocampus. This aligns with current models of cue reactivity in SUDs, which posit that drug cues activate circuits associated with reward processing, memory, and salience attribution(Cousijn et al., 2013; Goldstein & Volkow, 2002; Karoly et al., 2019; Koob & Volkow, 2010). Specifically, the frontal gyrus plays a crucial role in decision-making and impulse control, the insula contributes to interoceptive awareness and craving generation, and the hippocampus mediates memory consolidation and emotional processing (Everitt & Robbins, 2016; Rolls & Grabenhorst, 2008). These findings further support the notion that cue reactivity involves coordinated engagement of multiple brain regions underlying various aspects of addictive behavior.

### 4.3. Understanding Individual Variability in Cue Reactivity and Moderating Factors

While our study revealed overall trends in cue-elicited responses, the lack of robust relationships between neural and behavioral data in the latent variable space suggests significant individual variability in cue reactivity. This is consistent with growing evidence indicating individual differences in the neurobehavioral correlates of SUDs(Belin et al., 2008; Leggio et al., 2009). Future research should explore factors contributing to individual variability, such as **genetic predispositions, personality traits, environmental influences, and individual differences in reward sensitivity**. Additionally, it is crucial to explore **moderating factors** that might influence cue reactivity, such as current abstinence status, severity of dependence, and co-occurring psychiatric disorders (Ekhtiari et al., 2022). These investigations can further inform the development of personalized treatment approaches tailored to specific vulnerabilities and risk factors.

### 4.4. Limitations and Future Directions

We acknowledge the limitations of our study. Firstly, our participants were predominantly male, that could limit generalizability of results. Future investigations should explore potential sex differences in cue reactivity and include diverse samples considering age, ethnicity, and socioeconomic status. Secondly, relying on self-reported measures introduces potential biases. Future studies could incorporate objective physiological measures (e.g., heart rate, skin conductance), ecological momentary assessment methods (e.g., real-time craving reports), and implicit measures (e.g., implicit association tests) to enhance data sensitivity and ecological validity. Thirdly, our study lacked a control group of non-cannabis users. This limits our ability to definitively attribute the observed neural and behavioral responses to cannabis cue reactivity specifically. The observed differences could be due to pre-existing differences between cannabis users and non-users, rather than being directly caused by exposure to cannabis cues. Including a control group in future studies would allow for a more conclusive determination of the specific effects of cannabis cues on brain activity and subjective experience. Fourthly, although our study identified brain regions activated during cue exposure, further research is needed to elucidate the specific neurotransmitter pathways and cognitive processes mediating cue reactivity. This knowledge could inform the development of targeted interventions aimed at specific neural and cognitive mechanisms underlying relapse vulnerability.

### 4.5. Clinical Implications and Potential Interventions

Our findings hold significant implications for the development of evidence-based interventions for CUD. The observed individual variability in cue reactivity underscores the need for personalized treatment approaches. Such approaches could involve tailored cognitive-behavioral therapy (CBT) incorporating cue exposure therapy or mindfulness training focusing on individual vulnerabilities and reactivity patterns. Additionally, identifying specific brain regions and cognitive processes involved in cue reactivity could inform the development of targeted interventions such as:

- Neuromodulation techniques: Using transcranial magnetic stimulation (TMS) or transcranial electrical stimulation (tES) to modulate activity in specific brain regions implicated in cue reactivity.
- Pharmacological interventions: Developing medications targeting specific neurotransmitter pathways involved in reward processing and craving generation.
- Virtual reality exposure therapy: Utilizing VR technology to create immersive simulations of high-risk situations with cannabis cues, allowing individuals to practice coping skills in a safe and controlled environment.

Furthermore, understanding the triggers and mechanisms of cue reactivity can inform the development of preventative strategies, such as psychoeducational programs aimed at raising awareness about cue reactivity and teaching individuals coping skills to manage cravings in high-risk situations.

## 5. Conclusion

This study provides valuable insights into the neural and behavioral correlates of cannabis cue reactivity, as well as a pipeline for the cue validation process. We employed a multimethod approach to identify and validate cannabis cues capable of inducing craving and activating reward-related brain regions. Our findings highlight the role of individual variability and emphasize the need for personalized treatment approaches. By further exploring the specific mechanisms underlying cue reactivity and developing targeted interventions, future research can pave the way for more effective interventions and prevention strategies for CUD.

## Supporting information

Cue Validation Supplemental

## Ethics Statement

This study was carried out in accordance with the recommendations of the ethics board of the Iran University of Medical Sciences with written informed consent from all subjects. The protocol was approved by the ethics board of the Iran University of Medical Sciences.

## Acknowledgments

We would like to acknowledge the National Brain Mapping Lab (NBML) for providing invaluable support and assistance.

## Author Contributions

The concept of the study was developed by HE and TR. All authors contributed to the study design. ZH and PGh managed data collection, analyzed data, and wrote the first draft of the manuscript. PR and PGh ran the fMRI data analyses and contributed to the fMRI data interpretations. All authors participated in the revising of the manuscript.

## Data Availability

Publicly available datasets were analyzed in this study. This data can be found here: https://osf.io/85j6k/

